# Capability of GPT-4V(ision) in Japanese National Medical Licensing Examination

**DOI:** 10.1101/2023.11.07.23298133

**Authors:** Takahiro Nakao, Soichiro Miki, Yuta Nakamura, Tomohiro Kikuchi, Yukihiro Nomura, Shouhei Hanaoka, Takeharu Yoshikawa, Osamu Abe

## Abstract

**Background:** Previous research applying large language models (LLMs) to medicine was focused on text-based information. Recently, multimodal variants of LLMs acquired the capability of recognizing images.

**Objective:** To evaluate the capability of GPT-4V, a recent multimodal LLM developed by OpenAI, in recognizing images in the medical field by testing its capability to answer questions in the 117th Japanese National Medical Licensing Examination.

**Methods:** We focused on 108 questions that had one or more images as part of a question and presented GPT-4V with the same questions under two conditions: 1) with both the question text and associated image(s), and 2) with the question text only. We then compared the difference in accuracy between the two conditions using the exact McNemar’s test.

**Results:** Among the 108 questions with images, GPT-4V’s accuracy was 68% when presented with images and 72% when presented without images (*P* = .36).

**Conclusions:** The additional information from the images did not significantly improve the performance of GPT-4V in the Japanese Medical Licensing Examination.

## Introduction

The field of natural language processing is rapidly developing with the advent of large language models (LLMs). LLMs are models trained with massive text datasets and achieve the capability to understand and generate text in natural languages. With the introduction of ChatGPT [1] and other LLM-based chatbot services, many people have started to benefit from the use of LLMs. Although ChatGPT and its underlying model, Generative Pre-trained Transformer (GPT) [2,3], were not specifically developed for medical purposes, they possess a considerable amount of medical knowledge. They have achieved good scores in the United States Medical Licensing Examination [4] and are being explored for various applications for clinical and educational purposes [5–7]. GPT can also understand languages other than English. The latest model, GPT-4, has been reported to achieve passing scores in medical licensing examinations in non-English speaking countries such as Japan, China, Poland, and Peru [8–13].

Despite these successes, there is still a significant challenge in applying LLMs to real-world problems with non-text-based information. Radiological, pathological, and many other types of visual information play a crucial role in determining a patient’s management. Very recently, researchers have proposed multimodal variants of LLMs that can handle not only text but various types of input including images [14]. Providing medical images to multimodal LLMs may realize an even higher accuracy in solving medical-related problems. However, in previous studies on the accuracy rate of medical licensing examinations, questions with images were either not mentioned at all or explicitly excluded from the study. To the best of our knowledge, there is no study that directly evaluated the performance in solving questions with images. Therefore, in this study, we investigated the capabilities and limitations of GPT-4V [3,15], one of the most potent publicly available multimodal (vision and language) models, using the Japanese National Medical Licensing Examination as a subject.

## Methods

### Overview

From the questions of the 117th Japanese National Medical Licensing Examination, held in February 2023, we focused on those that included images as part of a question. Since some of these questions can be answered correctly without interpreting images, we measured the benefit of adding image information by comparing the accuracy rates of ChatGPT under two different conditions: 1) with both the question text and associated image(s), and 2) with the question text only.

### Dataset Details

Figure 1 shows the summary of our dataset. The questions and correct answers of the 117th Japanese National Medical Licensing Examination are publicly available for download on the official website of the Ministry of Health, Labor and Welfare [16]. All the questions are in a format in which a specified number of choices, typically one, are to be selected from five options. Of the questions that had images, two were officially excluded from scoring because they were either too difficult or inappropriate. Additionally, for two questions, images of female genitals were not made public on the aforementioned website. These four questions were excluded from our study.

**Figure 1.**
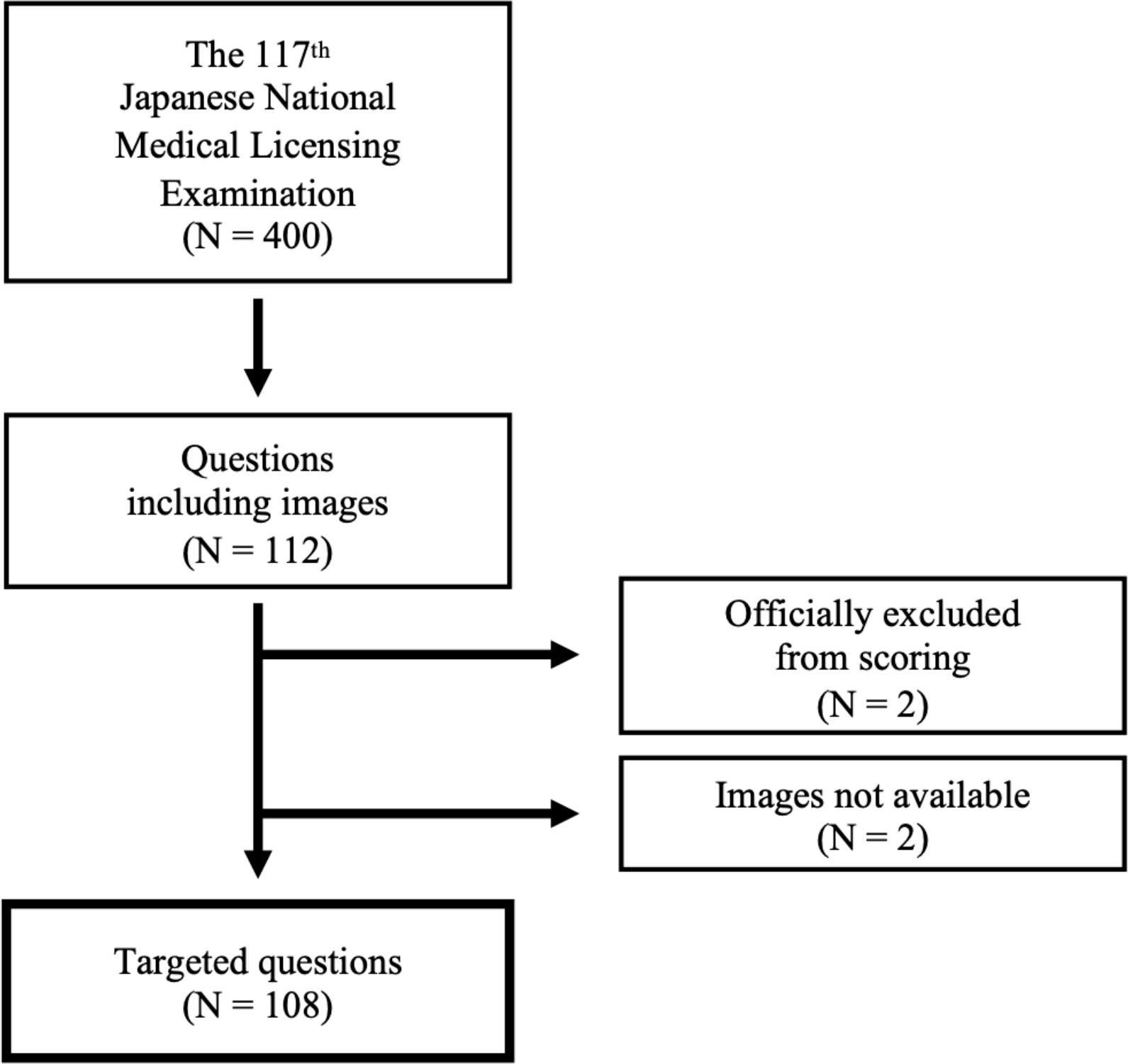
Summary of the questions included in this study.

The questions in the Japanese National Medical Licensing Examination were divided into two categories: clinical questions and general questions. In clinical questions, clinical information about a specific case is first presented, such as medical history and test results, and answers to questions about the case are required. General questions are about basic medical knowledge, and one is required to choose the correct answer among options for a short question text (typically of 1 or 2 sentences) with an image.

Some clinical questions consisted of multiple subquestions, in which case the background common to all the subquestions was first described, followed by the subquestions. In such cases, each subquestion was individually included in the following analysis if either the subquestion itself or the background part contained an image.

As a result, counting subquestions individually, out of 400 (sub-)questions, we collected 108 questions that had images, such as photographs of lesions, radiographic images, histopathological images, electrocardiograms, and graphs representing statistical data. Among them, 98 were clinical questions and 10 were general questions.

### Experimental Details

We used ChatGPT (September 25, 2023 version) enabled with GPT-4V, which is a multimodal model capable of processing both text and images. This version of ChatGPT asserts it was trained with information up to January 2022, meaning that it had no direct prior knowledge about our target examination. All the question statements and images were manually entered through ChatGPT’s web interface.

One of the authors, T.N., who has ten years of experience as a medical doctor, reviewed the outputs to interpret the response output by ChatGPT.

A new chat session was created for each question and each condition (i.e., with or without images). For questions that comprised multiple subquestions, the background information part and each subquestion were entered into ChatGPT in this order within the same chat session. Subquestions without images were also input to provide ChatGPT with enough context, but they were excluded from the accuracy calculations and the subsequent statistical analysis described below.

The questions were presented to ChatGPT without any preceding or custom instructions. Sometimes, ChatGPT did not respond with the specified number of choices, in which case an additional instruction, such as “Select only one option” or “Select two options”, was provided in Japanese. This additional instruction produced the correct number of options for all the questions.

### Statistical Analysis

The difference in ChatGPT’s performance between the two conditions (i.e., with or without images) was analyzed using the exact McNemar’s test. A *P*-value of less than .05 was considered statistically significant. The analysis was conducted using R (version 4.3.1; R Foundation for Statistical Computing, Vienna, Austria).

### Ethical Considerations

This study was conducted solely using publicly available resources; therefore, approval from the Institutional Review Board of our institution was not required.

## Results

Table 1 shows the results of our experiment. ChatGPT correctly answered 68% (73/108) of image-based questions when provided with both the question text and images, whereas it correctly answered 72% (78/108) of image-based questions when only the question text was provided. There was no significant difference in accuracy between these two conditions (*P* = .36). For the clinical questions, the accuracies when presented with and without images were 71% (70/98) and 78% (76/98), respectively. For the general questions, the accuracies were 30% (3/10) when presented with images and 20% (2/10) without images.

**Table 1.** Performance of ChatGPT in answering questions from the 117th Japanese National Medical Licensing Examination, when presented with or without associated images for each question.

| **Overall (P = .36)** |  | **With Images** |  |  |
| --- | --- | --- | --- | --- |
|  |  | **Correct** | **Incorrect** | **Total** |
| **Without Images** | **Correct** | 66 (61%) | 12 (11%) | 78 (72%) |
|  | **Incorrect** | 7 (6%) | 23 (21%) | 30 (28%) |
|  | **Total** | 73 (68%) | 35 (32%) | 108 (100%) |
| **Clinical (P = .21)** |  | **With Images** |  |  |
|  |  | **Correct** | **Incorrect** | **Total** |
| **Without Images** | **Correct** | 65 (66%) | 11 (11%) | 76 (78%) |
|  | **Incorrect** | 5 (5%) | 17 (17%) | 22 (22%) |
|  | **Total** | 70 (71%) | 28 (29%) | 98 (100%) |
| **General (P = 1.0)** |  | **With Images** |  |  |
|  |  | **Correct** | **Incorrect** | **Total** |
| **Without Images** | **Correct** | 1 (10%) | 1 (10%) | 2 (20%) |
|  | **Incorrect** | 2 (20%) | 6 (60%) | 8 (80%) |
|  | **Total** | 3 (30%) | 7 (70%) | 10 (100%) |

## Discussion

### Principal Results

In this study, we examined the image recognition capabilities of GPT-4V using questions associated with images from the Japanese National Medical Licensing Examination. To the best of our knowledge, this is the first study in which the capability of multimodal LLM for the Japanese National Medical Licensing Examination was investigated. Contrary to our initial expectations, the inclusion of image information did not result in any improvement in accuracy. Instead, we even observed a slight decrease, albeit not significant. This indicates that, at the moment, GPT-4V cannot effectively interpret images related to medicine.

For the clinical questions, in which sufficient clinical information including patient history was available in the text form, GPT-4V was able to choose the correct answers solely from the textual information in the majority (78%) of questions, but the addition of images did not improve the accuracy. On the other hand, for the general questions, there was little information in the question text, and GPT-4V had to determine the correct answer by interpreting the images. For these, GPT-4V yielded an accuracy rate that was hardly any better than random guessing even when presented with images. Our results suggest that, for both categories of questions, GPT-4V failed to utilize visual information to improve its accuracy. In our retrospective review, even among questions to which GPT-4V gave the correct answer only when presented with images (N=7), we found that GPT-4V often either did not mention the given image in its responses or gave an incorrect interpretation of it.

ChatGPT may serve as a valuable interactive teaching assistant in medical education; however, the inaccuracies in its responses are a significant concern [5,7]. Our current findings suggest that, especially with medical-related images, GPT-4V should not be relied upon as a primary source of information for medical education or practice. If used, extreme caution should be exercised regarding the accuracy of its responses. OpenAI officially states [15] that they “do not consider the current version of GPT-4V to be fit for performing any medical function or substituting professional medical advice, diagnosis, or treatment, or judgment” due to its imperfect performance in the medical domain. Yang et al. [17] have comprehensively examined the capabilities of GPT-4V in various tasks including medical image understanding and radiology report generation, and they stated that GPT-4V could correctly diagnose some medical images. However, as they acknowledge, their results contained a considerable number of errors, such as overlooking obvious lesions and errors in laterality. According to the case studies by Wu et al. [18], GPT-4V could recognize the modality and anatomy of medical images, but it could hardly make accurate diagnoses and its prediction heavily relied on the patient’s medical history. The results of our experiment supported these previous reports.

Considering the well-known high performance of GPT-4V in more generic image recognition tasks [3,17], the most probable reason for its limited image recognition performance in the medical field is that it may simply not have been trained with a sufficient number of medical-related images. LLMs are trained with a vast dataset available on the Internet, but medical images are not as readily accessible, partly due to privacy concerns. Some researchers are now working on developing multimodal LLMs specialized for medicine based on open-source LLMs [19,20] and using published or open resources. Moreover, although there are limited medical-related images publicly available on the Internet, hospitals have a vast amount of image data. A large part of which is accompanied by textual interpretations in the form of reports or medical records, which may serve as an ideal dataset for training multimodal LLMs. In highly specialized domains such as medicine, there remains a significant value in developing such domain-specific models.

### Limitations

This study had several limitations. Firstly, ChatGPT was not given any prior instructions and was directly presented with only the questions themselves. This might have negatively affected its capability to interpret images as the capabilities of LLMs are known to be affected by such “prompt engineering”. This will be a subject for future investigation. Secondly, the input was in Japanese. ChatGPT’s proficiency in non-English interpretation is known to be inferior to that In English interpretation. Translating the question text into English before inputting it to ChatGPT might have improved the model’s image interpretation capability.

However, in a previous study by Yanagita et al.[10], in which non-image questions from the Japanese National Medical Licensing Examination were the target, satisfactory results were achieved even when the questions were input in Japanese.

Thus, we adopted the same approach in our study. Thirdly, although our results were based on the same version of ChatGPT and the same question was evaluated with and without images on the same day, we cannot exclude the possibility that different models were used internally. Lastly, only a single evaluation was conducted for each condition and question. ChatGPT’s outputs have some randomness, and responses may differ across multiple evaluations. With ChatGPT’s application programming interface (API), users can programmatically control the degree of randomness by specifying a parameter called *temperature* and obtain mostly deterministic responses. However, during the time of this study, the API for GPT-4V was not available.

## Conclusions

At present, GPT-4V’s capability to interpret medical images may be insufficient. In highly specialized fields such as medicine, it is considered meaningful to develop field-specific multimodal models.

## Data Availability

All data produced in the present study are available upon reasonable request to the authors.

## Acknowledgements

The Department of Computational Radiology and Preventive Medicine, The University of Tokyo Hospital, is sponsored by HIMEDIC Inc. and Siemens Healthcare K.K.

## Abbreviations

LLM: large language model
GPT: generative pre-trained transformer
API: application programming interface

